# Dynamic programming model for pattern recognition on the *Pf*HRP2 sequence variants: a smart approach to improve malaria immunodiagnostics

**DOI:** 10.1101/2025.08.15.25333763

**Authors:** Ifeanyichukwu Okeke, Ukpe Ajima

## Abstract

The issue of poor diagnostic outcomes with malaria rapid diagnostic tests (mRDTs) is a serious concern. The World Health Organization (WHO) recommends confirming all suspected malaria cases through parasite-based diagnostic testing to ensure early detection, timely treatment, and minimize transmission risks. The mRDTs are crucial tools for this purpose and should consistently provide reliable results to ensure that patients receive appropriate treatment. In this study, a Python-based dynamic programming algorithm for recognizing unique patterns on genomic sequences was scripted and implemented to discover the presence, range, and frequency of epitope motifs in the genomic sequences of different *Pf*HRP2 isolates. Twenty three amino acid sequence variants of *Pf*HRP2 obtained from various databases were investigated. The investigations revealed that the majority of the unreviewed sequences had an abundance of different *Pf*HRP2 epitope motifs at multiple sites, whereas these motifs were scarce among the reviewed sequences.

## INTRODUCTION

### Background to the Study

*Plasmodium falciparum* histidine-rich protein 2 (*Pf*HRP2) is an important biomarker for *Plasmodium falciparum* (*Pf*), the primary cause of severe malaria in humans, especially in tropical regions. To contain malaria and reduce morbidity and mortality arising from it, there is a need for accuracy in its diagnosis. A lot of progress has been made so far towards the elimination of malaria. A particular reference is a targeted initiative of the World Health Organization (WHO) – “high burden to high impact (HBHI)” aimed at accelerating progress against malaria. Following this intervention, mortality rates in eleven African countries have dropped by 13% between 2017 and 2023. Since 2015, many countries, including Azerbaijan, Belize, Cabo Verde, Egypt, and Tajikistan, and recently over the past two years, nine other ones have been certified as malaria-free by the WHO [1]. These progress reports are expected considering demonstrated commitments and investments towards malaria elimination by the global body.

The flexibility of *Pf* to alter its genetic make-up and potentially change what is known about its immunodiagnostics, where immune epitopes become altered by deletion, single amino acid substitution, or addition, or emergence of new ones. The alteration of the immune epitopes poses the challenge of generating new antigens different from what is originally in use for the detection of the parasite. This phenomenon is a major reason for poor performance often seen with the malaria rapid diagnostic tests (mRDTs), where a test kit produces a false negative result following antigen – antibody mismatch apart from other factors such as protein folding, antigen concentration, genomic instability, HRP2 gene deletions, low parasite density, improper use and storage etc.

In this study, we focused on the antigen *Pf*HRP2. It is the most abundant antigen carried by the *Pf* species; hence, its detection is a specific diagnosis for *Pf*. There have been several reports of poor performance and inconsistent results surrounding the mRDT from users as reported by [2]. For this reason, we seek to explore the *Pf* immunodiagnostic epitopes about the diversity of *Pf*HRP2 from different isolates, as a means of boosting the sensitivity of *Pf* rapid diagnostic testing.

### Dynamic programming and *Pf*HRP2 sequence variants analysis

Dynamic programming (DP) is an algorithmic technique applied in computational biology, particularly in genomic sequence analysis. It is an essential tool in genomic data analysis, facilitating the efficient and accurate alignment of sequence data. *Pf*HRP2, whose gene exhibits significant sequence variation, is a key diagnostic target for P. falciparum malaria. Sequence variations that particularly occur in the repeat regions are the leading cause of false-negative results in RDTs. DP plays a crucial role in analyzing the different genomic patterns in these variants [3, 4].

DP breaks down a complex problem into smaller overlapping subproblems. The solutions to these subproblems are then stored and reused, avoiding redundant calculations and drastically improving computational efficiency [5]. The use of DP in the analysis of *Pf*HRP2 sequence variants highlights the importance of computational approaches in understanding genetic diversity and its implications for disease diagnosis and treatment. DP offers a valuable tool for unraveling the complex relationships between genetic variation and disease phenotype, enabling more effective malaria control strategies and improved diagnostic tools [6].

In this study, we use DP to align *Pf*HRP2 sequence variants from different parasite isolates, identifying the presence/absence, positions, and frequency of occurrence of identified epitope motifs. This information is useful in understanding the impacts of these epitopes on mRDT performance. The information can also provide valuable insights into the evolution, distribution, diagnostic implications of this important malaria protein, as well as the effects of mutation on protein folding and stability.

### Genomic sequence variation and mRDT sensitivity

Different authors have different opinions regarding genomic sequence variation and the sensitivity of mRDT. While some studies [7] & [8] suggest that sequence variation does not affect mRDT sensitivity, [9] suggested that sequence variation is associated with RDT nonsensitivity. Many other authors [10, 11, 12, 13] believe that the correlation between RDT performance and *Pf*HRP2 sequence variation remains unclear. It has been suggested that alterations in the amino acid composition of this protein could be the reason for reduced RDT sensitivity, and issues of this nature may be a factor responsible for the low positive rate and low accuracy often observed with the mRDTs.

Previous reports [7] have indicated that deletion of *Pf*HRP2 in the Pf genome affects the accuracy of *Pf*HRP2-based RDT. However, [13] and [8] agreed that despite extreme variation, the diversity of *Pf*HRP2 does not appear to affect RDT performance but only nPCR. In addition to antigenic diversity, other factors, such as persistent antigenaemia and parasite density, also affect the accuracy of *Pf*HRP2-based test kits [14].

Expanding the parasite detection spectrum by applying multiple MoAbs on a detection kit will be a game changer in malaria rapid detection testing, also serving as a turning point in the argument surrounding the effect of sequence variations on mRDT sensitivity. In this study, we examined the presence of already known epitopes of *Pf*HRP2, their positions, and frequencies on the sequences of selected variants. The abundance of these immunogenic epitopes on the *Pf*HRP2 sequences of several variants could be harnessed in the synthesis of more potent specific antibodies that could expand the detection spectrum of *Pf* using the RDT kits.

It is not to our knowledge how often mRDT kit manufacturers review the viability of their custom antibodies in detecting new and emerging malaria strains. We supposed that they use antibodies derived from the epitopes of the reviewed sequences, which are no longer up to date, as newer, unreviewed sequences tend to harbour more of the epitopes identified from sequences from recent studies. Except that kit manufacturers are using antibodies derived from epitopes from the NCBI, those from UniProt-reviewed sequences tend to lack sufficient epitope diversity and concentration that can enhance diagnostic sensitivity. Some sequences in this group have a complete absence of any of the identified epitopes (Table 1).

This whole study was done *in silico*; no in vitro laboratory procedure was involved. In silico techniques, though, are predictive, but are also valid and acceptable means in scientific discovery. It is a useful technique in the fields of biotechnology and drug discovery, where it is applied to reduce costs and the time required to complete a procedure manually. Though not much work has been done in the aspect of *Pf*HRP2 variation on mRDT performance via in silico, a good number of works have been done even in the most critical aspect of drug discovery and vaccine development (immunoinformatics) [9, 10, 11, 12, 15, 16, 17, 18].

MRDTs are quick and portable immunochromatographic lateral flow devices that rely on the capture of parasite antigens by monoclonal antibodies. These diagnostic antigens result from repetitive sequences that can form through slipped-strand mispairing during DNA replication or unequal crossover of chromosomes during meiosis [19]. Generally, a rapid diagnostic test is a medical diagnostic test that is quick and easy to perform. RDTs are suitable for preliminary or emergency medical screening and use in medical facilities with limited resources. They also allow point-of-care testing in primary care for things that formerly only a laboratory test could measure. They provide same-day results within two hours, typically in approximately 20 minutes [20, 21].

### Epitopes of the *Pf*HRP2 sequences as diagnostic biomarkers for *Plasmodium falciparum* malaria

There are several antigenic epitope repeat patterns in the *Pf*HRP2 molecule to which its monoclonal antibodies in the *Pf*HRP2-based rapid diagnostic test kit can bind. Some of these patterns have been listed here. As observed from this study, the reviewed sequences from the UniProt, which are considered to have been curated, did not display as much epitopes as expected. The highest number of epitopes observed in this study was 22, which was displayed by tr|Q3ZJI4|Q3ZJI4_PLAFA. Apart from sp|P05227|HRP1_PLAFA and a few other sequences that had up to 18 epitopes, sp|P04930|HRP_PLAFF and sp|P05228|HRP2_PLAFA never had more than two. All the unreviewed sequences except tr|Q8IDG8|Q8IDG8_PLAF7 and all four sequences from the NCBI database have abundant epitopes.

Only three out of the ten of the reviewed sequences investigated had epitope motifs in their sequences. The rest of the isolates of the reviewed sequences will evade detection by mRDT utilizing any or a combination of the available epitopes in mRDT antibody formulation.

A major concern is what happens when AAYAHHAHHAAY is being utilized to formulate the antibodies in the mRDT kits in use. Here lies the reason why mRDT kit manufacturers should disclose the epitope and antibodies in their products.

### Salient factors affecting the accuracy and sensitivity of malaria rapid diagnostic testing

Apart from antigen-antibody mismatch, incorrect folding of the 3D protein shape of the antibody on the diagnostic disc is another major cause of false-positive and false-negative malaria test results with the RDT kits. Incorrect folding of the 3D protein shape is a major cause of mRDT insensitivity. In the event of compromised shape, the antigen fails to bind to the antibody even when the parasite is present, or the antibody binds to some sites where it shouldn’t, causing false-positive or false-negative results. While false positives are less common, the more common false negative is a dangerous outcome.

Incorrect protein folding could arise from environmental factors, such as storage conditions. Extremes of temperature, pH, and humidity disrupt protein folding. Antibodies have optimal temperature, pH, and humidity at which they are most effective, as deviation can affect their folding. Handling of antibodies during the production and purification processes can also impact their folded structure. Other factors, such as antigen concentration, genomic instability, HRP2 gene deletions, low parasite density, and improper use, also contribute to poor RDT performance given an exact antigen-antibody match.

## MATERIALS AND METHODS

### Materials

This in silico study was performed via a computer set with the following applications and software: Python 3.13.2 integrated development environment (IDE), Google Chrome internet browser, good internet connectivity, Internet Download Manager (IDM), the UniProt and NCBI databases. This work was carried out on Windows 11 Pro with Processor: Intel(R) Core (TM) i5 CPU M 520 @ 2.40GHz, 2400Mhz, 2 Core (s), 4 Logical Processor (s).

### Methods

#### *Pf*HRP2 genomic sequences and antigenic epitopes mining

The general information and amino acid sequence variants of *Pf*HRP2 were extracted from UniProt [22] and NCBI [23]. Data mining of antigenic epitopes was performed through a literature search. A DP algorithm for epitope motifs presence, location (range), and counts (frequency of occurrence) on the sequence variants was scripted and implemented on the Python IDE as described below.

The naïve approach to search for the occurrence of a pattern *p* (an epitope in this situation) of length *k* in a sequence *s* of length (N > k) considers all possible sub sequences of *s*, of size *k*, and compares each of those, position by position, to *p*. Once a mismatch is found, we move on to test the next sub sequence. If the subsequence matches the pattern in all positions, an occurrence of *p* is found. If the purpose is to find all occurrences of *p* in *s*, we continue even in case of success (Figure 1), while if it is only required to find the first occurrence (or simply if the epitope occurs in the sequence), we can stop when the first sub sequence matches *p* (Figure 2). the function returns the position of the first occurrence of the epitope, or if the epitope does not occur returns −1.

**Figure 1.**
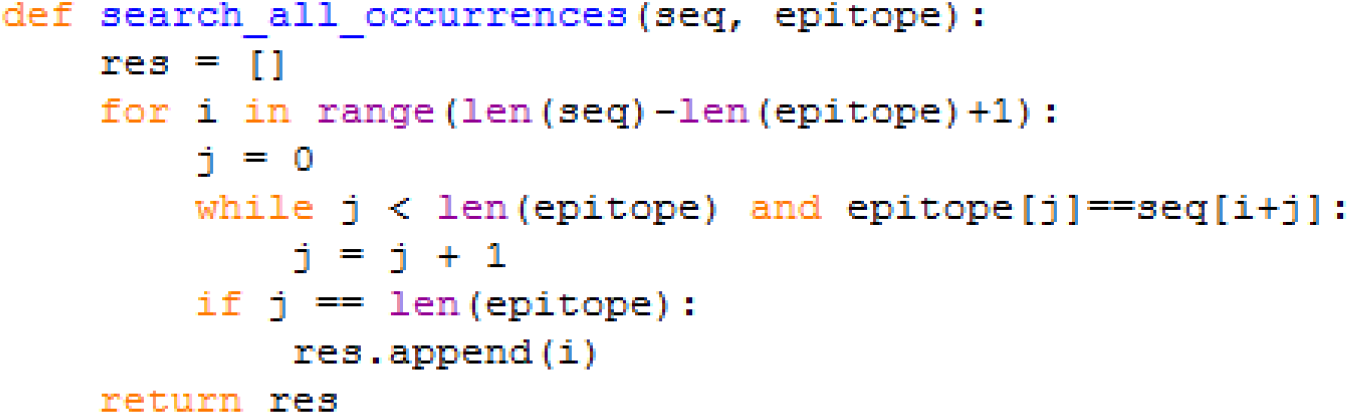
To search all occurrences of an epitope in a sequence.

**Figure 2.**
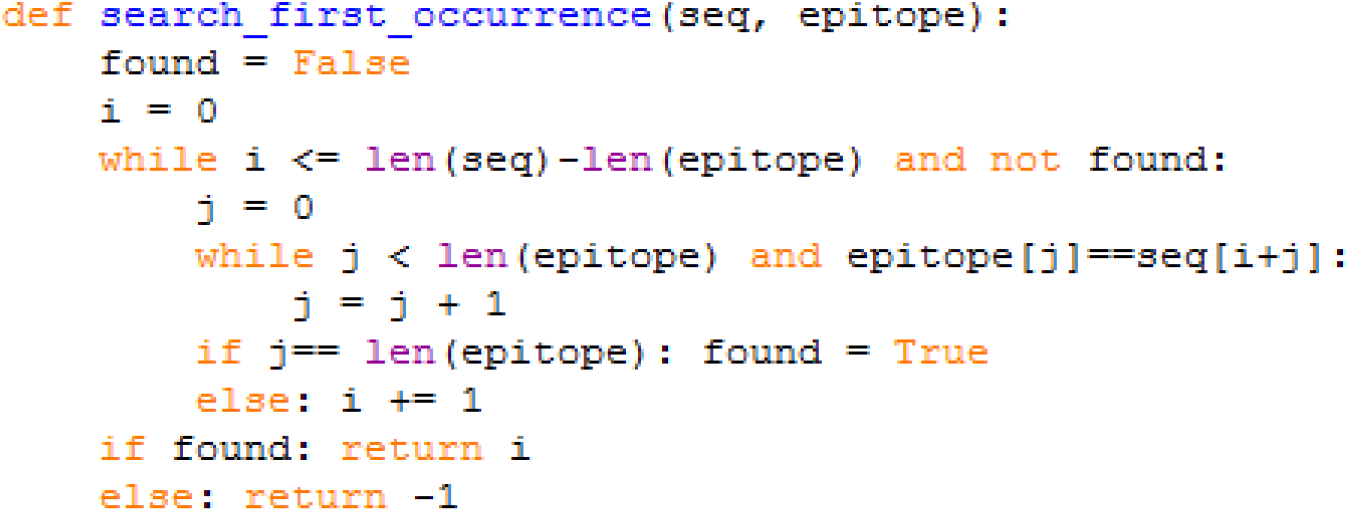
To search for the first occurrence of an epitope in a sequence.

The following code block was scripted to determines the range of epitope motifs on the sequences (figure 3).

**Figure 3.**
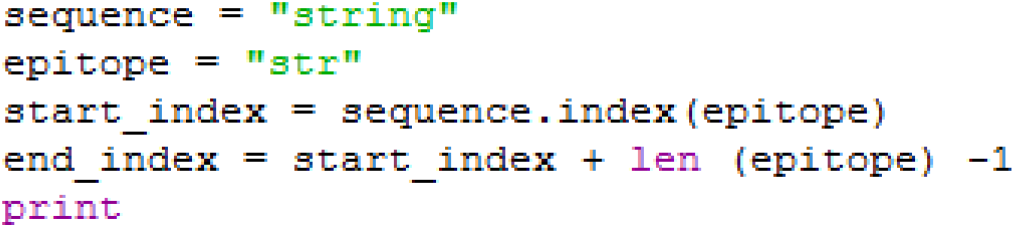
To determine the range of an epitope in a sequence.

Applying the codes above, by substituting the sequences and epitope motifs one after the other, the presence, frequency, and ranges of the epitope motifs on the sequences were identified. The results are provided in Table 1 as supplementary material B. Table 2 relates the frequency distribution of epitopes among the *Pf* genome. The results are presented as follows.

**Table 2.**
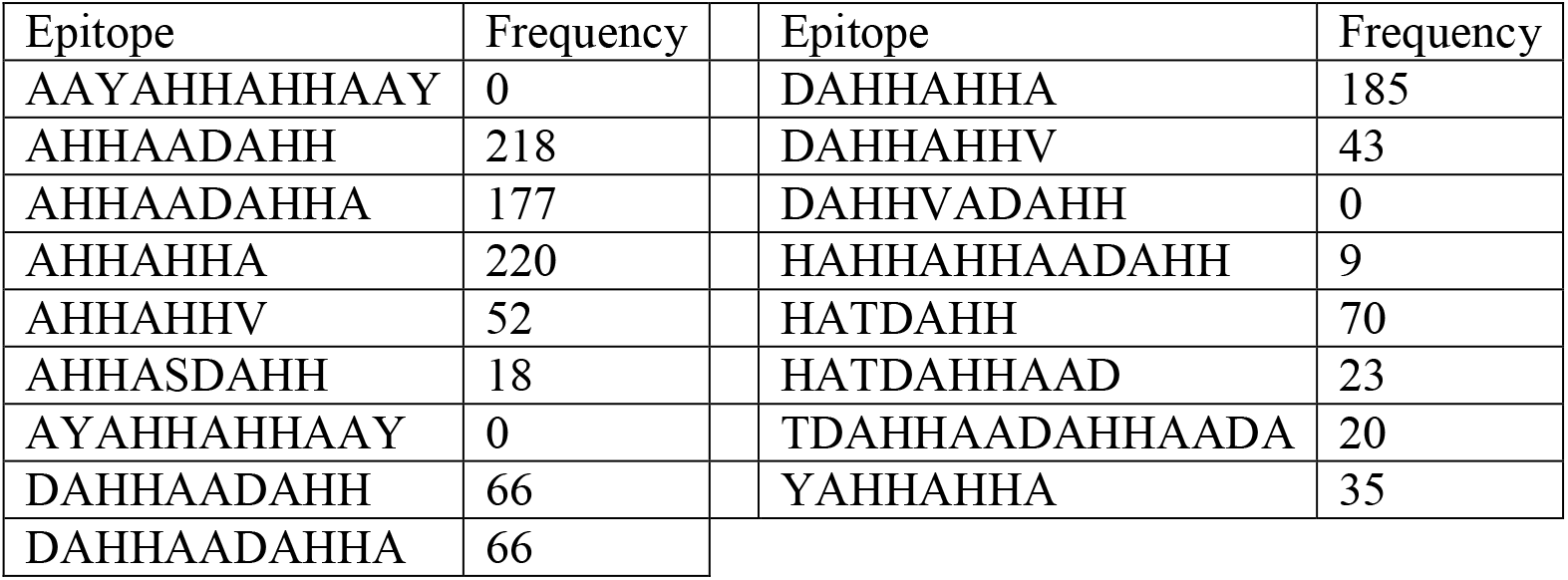
Frequency distribution of epitopes among the *Pf* genome.

## RESULTS

### Result Interpretation

A comprehensive list of the sequences of the isolates investigated in this study is presented in the list of sequences as supplementary material A. The study involved searching for the presence, location (range), and counts of the various antigenic epitopes of *Pf*HRP2 on the sequences of the selected isolates. Figure 4 is a Python code block returning the absence (0) and abundance of epitopes in two different *Plasmodium falciparum* isolates. Figure 5 is another code block showing a programme for determining the range of the epitopes within the sequences.

**Figure 4.**
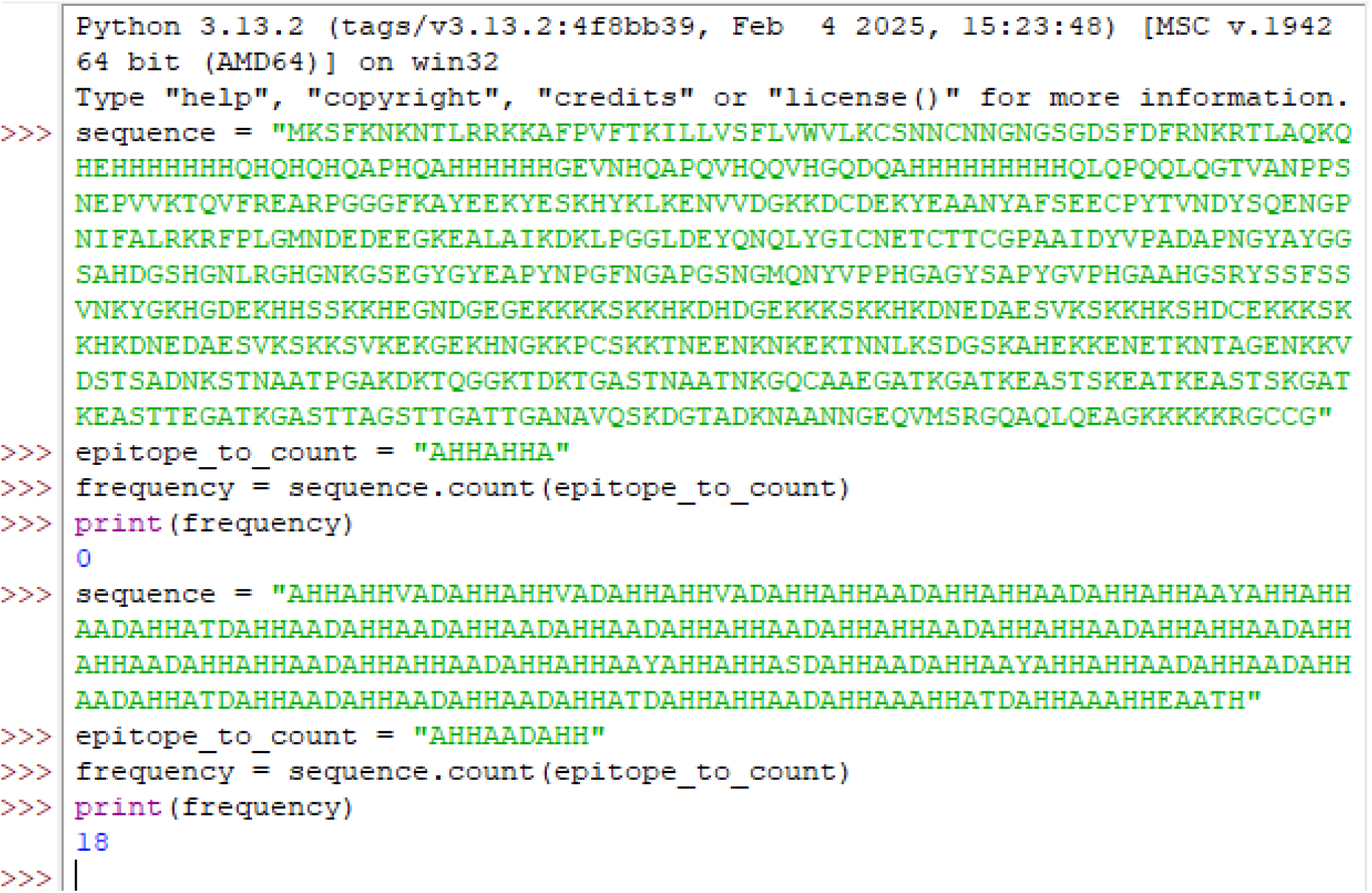
Programme determining the presence and frequency of epitope motifs on the sequences.

**Figure 5.**
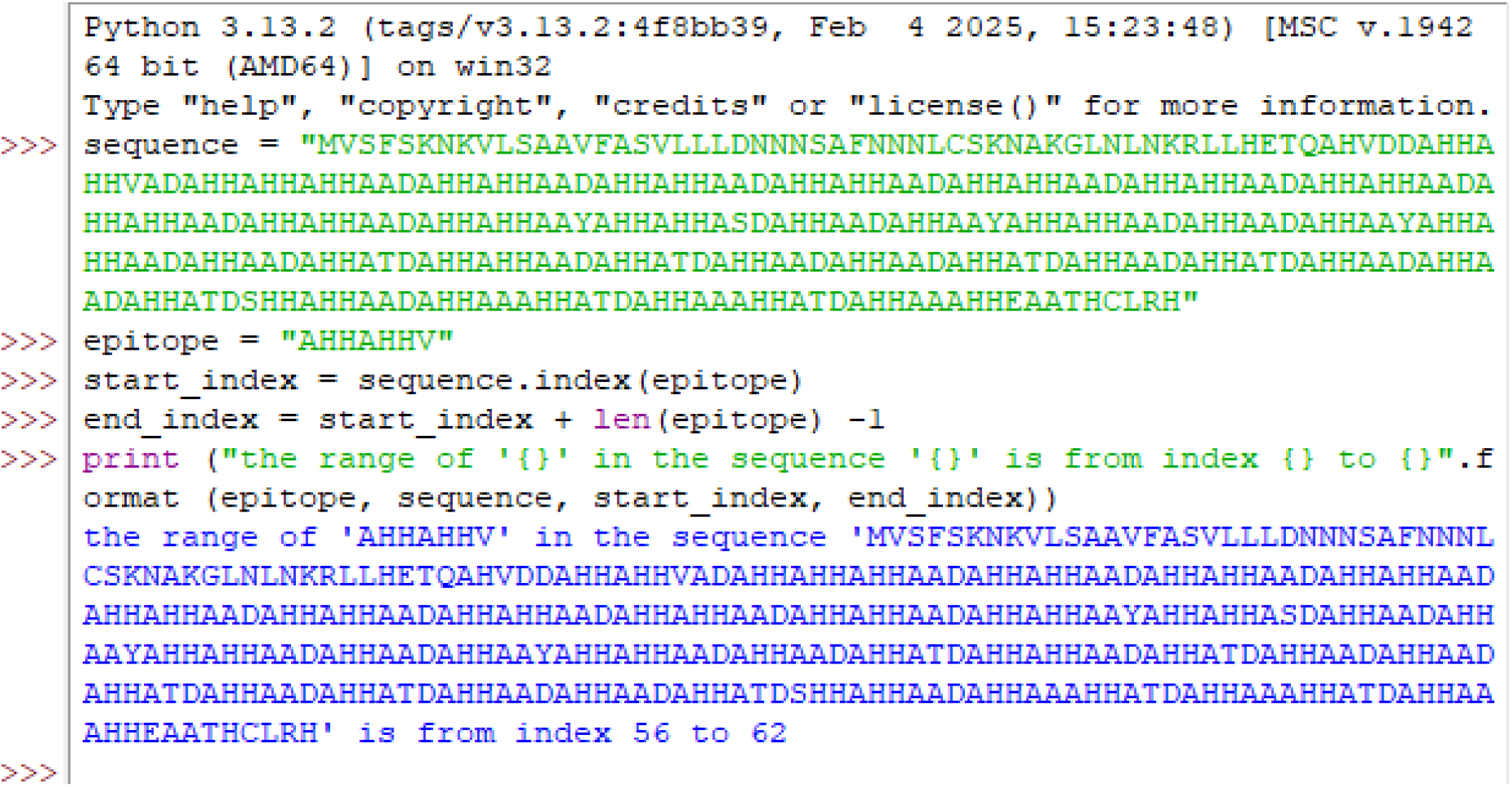
The programme determines the range of epitope motifs on the sequences.

### Sequences of *Pf*HRP2 used in the Study

A total of 21 *Pf*HRP2 sequence variants were retrieved from the UniProt database. Among these variants, the first 8 numbered 1-8 were reviewed sequences (Swiss-Prot), and the next eleven numbered 1a-1k were unreviewed (TreEMBL). An additional four sequences numbered 22-25 were extracted from the NCBI database (supplementary material A). Biological sequence databases are classified into two categories: primary and secondary databases. The primary databases contain sequence data submitted by the researchers, not fully processed and not curated. The major primary databases include ENI / EMBL-bank (www.ebi.ac.uk/ena), GenBank (or UniProt (www.uniprot.org/uniprot, www.ncbi.nlm.nih.gov/GenBank)), DDBJ (www.ddbj.nig.ac.jp), and RefSeq. Secondary databases contain data obtained from primary databases and have been curated by specialists in terms of consistency and completeness. An example is the NCBI database.

### Epitope search

The epitopes used in this study were those listed in previous studies by [24, 25, 26]. A compilation of the epitopes is listed as follows: AAYAHHAHHAAY, AHHAADAHH, AHHAADAHHA, AHHAHHA, AHHAHHV, AHHASDAHH, AYAHHAHHAAY, DAHHAADAHH, DAHHAADAHHA, DAHHAHHA, DAHHAHHV, DAHHVADAHH, HAHHAHHAADAHH, HATDAHH, HATDAHHAAD, TDAHHAADAHHAADA, and YAHHAHHA. These epitopes are the parts of *Pf*HRP2 that bind to monoclonal antibodies (MoAbs) incorporated onto the malaria rapid diagnostic test strip. A total of 17 epitopes were investigated. For DAHHAHHA, there could be substitutions of Y or V for the first and last amino acids, YAHHAHHA allows possible substitutions of D or V for its first or last amino acids, and DAHHAADAHH can substitute H and V at the first and fifth amino acids, respectively [27].

## DISCUSSION

Only three isolates (sp|P04930|HRP_PLAFF Small histidine-alanine-rich protein OS=*Plasmodium falciparum* (isolate FC27 / Papua New Guinea) OX=5837 PE=2 SV=1, sp|P05227|HRP1_PLAFA Histidine-rich protein *PF*HRP-II OS=*Plasmodium falciparum* OX=5833 PE=2 SV=1, and sp|P05228|HRP2_PLAFA Histidine-rich protein *PF*HRP-III OS=*Plasmodium falciparum* OX=5833 PE=2 SV=1) out of the eight of the reviewed sequences from the UniProt investigated had epitope motifs. This implies that only 37.5% of them possess the tendency to be detected using the immunodiagnostic method. It is also important to note that the majority of these isolates have low epitope concentrations as low as 5.9% and this significantly affects RDT sensitivity.

Sp|P05227|HRP1_PLAFA histidine-rich protein *PF*HRP-II OS=*Plasmodium falciparum* OX=5833 PE=2 SV=1 had all the epitope motifs represented in its sequence except AAYAHHAHHAAY, AYAHHAHHAAY, and DAHHVADAHH, also possessing as high as 100% epitope concentration with regard to AHHAADAHH. An abundance of epitopes in this isolate implies that it can be easily detected with most of the antibodies, except those formulated from the deficient epitopes. The rest of the reviewed sequences had none of the epitopes. This is a signal for possible false negative results should these isolates be tested with kits bearing antibodies from the listed epitopes. AHHAHHA has the highest occurrence among the sequences of the *Pf* genome; therefore, it could be a better option during epitope selection for *Pf* immunodetection. In contrast, AAYAHHAHHAAY, DAHHVADAHH, and AYAHHAHHAAY had no occurrences among the *Pf* genome; therefore, they could be a potential cause of false negative results and should be avoided (Figure 6).

**Figure 6.**
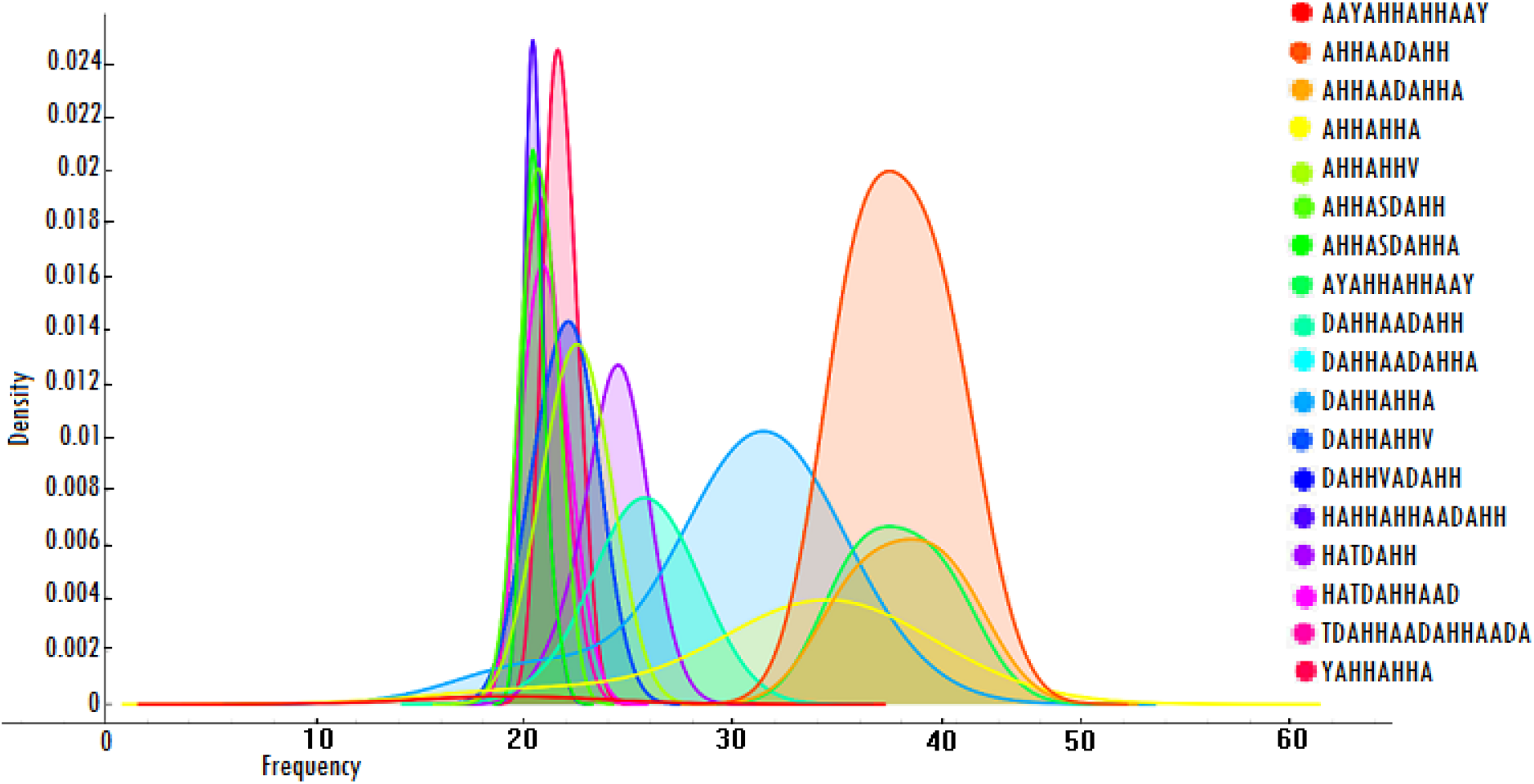
Epitope density among the *Pf* genome.

### Epitope motif prevalence among *Pf* isolates

The epitope AHHAHHA has the highest frequency of 220, followed by AHHAADAHH 218, making them epitopes of choice in monoclonal antibody formulation for *Pf* immunodiagnosis. On the other hand, AAYAHHAHHAAY, AYAHHAHHAAY, and DAHHVADAHH are not options at all when looking for diagnostic antibodies for *Pf*, as they do not occur in any of the sequences investigated, Table 1 and Figure 5. We want to believe that those epitopes with zero frequencies were not used in formulating any of the diagnostic antibodies in the mRDT kits.

### Genomic Sequence Variation of *Pf*HRP2 and Malaria RDT Sensitivity

A subtle point of inquisitiveness arose when we discovered that two 3D7 isolates (CZT62760.1 histidine-rich protein II [*Plasmodium falciparum* 3D7] and XP_002808743.1 histidine-rich protein II [*Plasmodium falciparum* 3D7]) from the NCBI database showed sequence dissimilarity with the unreviewed 3D7 isolate (tr|Q8IDG8|Q8IDG8_PLAF7 Membrane associated histidine-rich protein 2 OS=*Plasmodium falciparum* (isolate 3D7) OX=36329 GN=*PF*3D7_1353200 PE=4 SV=1) from the UniProt database. While those from the NCBI had a total length of 305 amino acids, those from UniProt had 137. In situations like this, there is bound to be a loss of accuracy due to the use of extraneous functional motifs when diagnostic or therapeutic formulations are made from information from either sequence.

*Pf*HRP2 is a biomarker specific for *Plasmodium falciparum*. Analysis of the amino acid sequence variants of *Pf*HRP2 provides significant insights into how protein antigenic diversity can affect the performance of the *Pf*HRP2-based rapid diagnostic test (RDT) kit and the reliability of the results generated via the *Pf*HRP2-based biomarker. The var genes [14, 29] of the parasite are constantly changing the expressible proteins of the parasite at every moment [30] and the diagnostic antigens are not shielded from being affected. It is therefore possible that the sequences initially used for the research may have altered drastically over time, which may adversely impact the efficiency of the antigen as a conventional diagnostic marker.

It has been reported that certain polymorphisms in the *Pf*HRP2 sequence, particularly when certain amino acid repeats are involved, do affect mRDT sensitivity [31]. These polymorphisms have been detected in the Asia-Pacific region, India, Madagascar [32, 33], and Uganda [34]. Additionally, gene deletion involving *Pf*HRP2 and *Pf*HRP3 also results in false-negative results by mRDTs. This has been reported in Peru [34, 35], Mali [36], India [37], and Brazil [38]. Variation in the amino acid repeat of *Pf*HRP2 can affect mRDT sensitivity, especially when such repeats are those involved in diagnostic prediction. The more diagnostic predicting repeats available in the sequence of *Pf*HRP2, the higher the sensitivity of the RDT model. In other words, the sensitivity of the RDT model is directly proportional to the antigen concentration.

### Summary and Conclusion

Our investigation of *Pf*HRP2 sequences of isolates obtained from the UniProt and NCBI databases indicated the presence of different epitope motif sites, mostly in the unreviewed sequences from the UniProt and NCBI isolates, while only a handful of the reviewed sequences contain epitopes. This implies that there is enough epitope concentration among a wide range of variants to improve the detection spectrum of mRDT products and enhance their sensitivity. An abundant concentration of epitopes in an antigen implies that, despite gene mutations that may occur in the parasite, unaffected epitope sites can still interact with the corresponding monoclonal antibodies used in the test kits, maintaining their effectiveness. Epitopes with the highest frequency per isolate and occurring in more variants should be mostly favoured in RDT formulation. It is important to note that large-scale mutations involving the epitope sites and the use of non-corresponding monoclonal antibodies (MoAbs) could significantly impact test sensitivity. Therefore, incorporating as many MoAbs as possible into the kits could greatly improve mRDT sensitivity.

### Limitations of the study and suggestions for future studies to solve the challenge

The major limitation of this study is the use of sequenced data from designated databases whose raw information is not within our disposal. Also, the epitope motifs used in this study were those mined from previous reports; the raw data are not within our reach. The implication of this is that the finding may not be a true reflection of the reality in the event of any alteration in the *Pf*HRP2 gene in a given region. For this reason, we propose further studies that pair pattern identification with sequenced data from isolates sourced from diverse geographic settings. This is necessary due to the important role mRDTs play in malaria control.

### Recommendation

Malaria diagnostic kit manufacturers do not disclose which of the epitopes is targeted in their products. We learned from interaction that manufacturers use recombinant constructs of the epitope’s corresponding antibodies in the mRDT kits rather than naturally produced antibodies. Indicating the epitope motifs whose antibodies were incorporated in the formulation of mRDT kits will play a critical role in the appropriate distribution of kits, as the use of an epitope in a region where the bearing isolate is not prevalent may lead to poor diagnostic outcomes with the mRDTs. Periodic epidemiologic and genomic surveillance studies should be conducted to monitor *Pf*HRP2 gene alterations capable of impacting its detection. We also recommend further studies to map immune epitopes on those reviewed sequences that do not exhibit any immune epitopes to enhance their detection using the mRDTs.

## Data Availability

All data produced in the present work are contained in the manuscript

## List of abbreviations

DP: Dynamic programming
MoAb: Monoclonal antibodies
mRDT: malaria Rapid Diagnostic Test
NCBI: National Center for Biotechnology Information
*Pf*: *Plasmodium falciparum*
*Pf*HRP2: *Plasmodium falciparum* histidine-rich protein 2
RDT: Rapid Diagnostic Test
WHO: World Health Organization

## Authors’ contributions

UA and IO conceptualized the study, IO carried out the investigations and analyses of datasets, and IO composed the manuscript, whereas UA supervised the entire research.

## Acknowledgements

We appreciate the management of the Africa Centre of Excellence in Phytomedicine Research and Development (ACEPRD), University of Jos, for providing the enabling platform for the study of bioinformatics and genomics at the university.

## Description of materials of this study

Supplementary material A: list of sequences of *Pf*HRP2 investigated in this study.

Supplementary material B: Table 1. Summary of epitope frequency among *Plasmodium falciparum* isolates.

