## Supplemental file A for "Dynamic programming model for pattern recognition on the *Pf*HRP2 sequence variants: a smart approach to improve malaria immunodiagnostics"

**Supplementary material A: Sequences of *Pf*HRP2 isolates used in the study**

NB: The reviewed sequences (S/N:1 – S/N:10) and the unreviewed sequences (S/N:1a – S/N: 1k) were retrieved from the UniProt database. The rest (S/N:22 – S/N:25) were retrieved from the NCBI database.

**Reviewed sequences**

>S/N:1

>Identifier: sp|P09346|KNOB_PLAFG Knob-associated histidine-rich protein OS=*Plasmodium falciparum* (isolate FCR-3 / Gambia) OX=5838 PE=2 SV=1

>Organism: *Plasmodium falciparum (isolate FCR-3 / Gambia)*

MKSFKNKNTLRRKKAFPVFTKILLVSFLVWVLKCSNNCNNGNGSGDSFDFRNKRTLAQKQ 60

HEHHHHHHHQHQHQHQAPHQAHHHHHHGEVNHQAPQVHQQVHGQDQAHHHHHHHHHQLQP 120

QQLQGTVANPPSNEPVVKTQVFREARPGGGFKAYEEKYESKHYKLKENVVDGKKDCDEKY 180

EAANYAFSEECPYTVNDYSQENGPNIFALRKRFPLGMNDEDEEGKEALAIKDKLPGGLDE 240

YQNQLYGICNETCTTCGPAAIDYVPADAPNGYAYGGSAHDGSHGNLRGHGNKGSEGYGYE 300

APYNPGFNGAPGSNGMQNYVPPHGAGYSAPYGVPHGAAHGSRYSSFSSVNKYGKHGDEKH 360

HSSKKHEGNDGEGEKKKKSKKHKDHDGEKKKSKKHKDNEDAESVKSKKHKSHDCEKKKSK 420

KHKDNEDAESVKSKKSVKEKGEKHNGKKPCSKKTNEENKNKEKTNNLKSDGSKAHEKKEN 480

ETKNTAGENKKVDSTSADNKSTNAATPGAKDKTQGGKTDKTGASTNAATNKGQCAAEGAT 540

KGATKEASTSKEATKEASTSKGATKEASTTEGATKGASTTAGSTTGATTGANAVQSKDGT 600

ADKNAANNGEQVMSRGQAQLQEAGKKKKKRGCCG 634

>S/N:2

>Identifier: sp|P04930|HRP_PLAFF Small histidine-alanine-rich protein OS=*Plasmodium falciparum* (isolate FC27 / Papua New Guinea) OX=5837 PE=2 SV=1

>Organism: *Plasmodium falciparum (isolate FC27 / Papua New Guinea)*

MVSFSKNKILSAAVFASVLLLDNNNSEFNNNLFSKNAKGLNSNKRLLHESQAHAGDAHHA 60

HHVADAHHAHHAANAHHAANAHHAANAHHAANAHHAANAHHAANAHHAANAHHAANAHHA 120

ANAHHAANAHHAANAHHAANAHHAADANHGFHFNLHDNNSHTLHHAKANACFDDSHHDDA 180

HHDGAHHDDAHHDGAHHDGAHHDGAHHDGAHHNATTHHLHH 221

>S/N:3

>Identifier: sp|P05227|HRP1_PLAFA Histidine-rich protein PFHRP-II OS=*Plasmodium falciparum* OX=5833 PE=2 SV=1

>Organism: *Plasmodium falciparum*

MVSFSKNKVLSAAVFASVLLLDNNNSAFNNNLCSKNAKGLNLNKRLLHETQAHVDDAHHA 60

HHVADAHHAHHAHHAADAHHAHHAADAHHAHHAADAHHAHHAADAHHAHHAADAHHAHHA 120

ADAHHAHHAADAHHAHHAADAHHAHHAADAHHAHHAAYAHHAHHASDAHHAADAHHAAYA 180

HHAHHAADAHHAADAHHAAYAHHAHHAADAHHAADAHHATDAHHAHHAADAHHATDAHHA 240

ADAHHAADAHHATDAHHAADAHHATDAHHAADAHHAADAHHATDSHHAHHAADAHHAAAH 300

HATDAHHAAAHHATDAHHAAAHHEAATHCLRH 332

>S/N:4
>Identifier: sp|P05228|HRP2_PLAFA Histidine-rich protein PFHRP-III OS=*Plasmodium falciparum* OX=5833 PE=2 SV=1
>Organism: *Plasmodium falciparum*

MVSFSKNKVLSAAVFASVLLLDNNNSEFNNNLFSKNAKGLNSNKRLLHESQAHAGDAHHA 60

HHVADAHHAHHVADAHHAHHAANAHHAANAHHAANAHHAANAHHAANAHHAANAHHAANA 120

HHAANAHHAANAHHAANAHHAANAHHAANAHHAANAHHAADANHGFHFNLHDNNSHTLHH 180

AKANACFDDSHHDDAHHDGAHHDDAHHDGAHHDDAHHDGAHHDGAHHDGAHHNATTHHLH 240

H 241

>S/N:5
>Identifier: sp|P14586|HRP3_PLAFS Histidine-rich protein OS=*Plasmodium falciparum* (isolate fcm17 / Senegal) OX=5845 PE=4 SV=1
>Organism: *Plasmodium falciparum (isolate fcm17 / Senegal)*

MLNHRYHHYFHRHHHLNHHLYHRHHHHRHHHHRHHHRHQILHQNRHQIHQILLSLNNKIM 60

GYAIFLFLSILLHLVYLVIHRL 82

>S/N:6
>Identifier: sp|P14587|YDH1_PLAFS Uncharacterized protein 5' to Asp-rich and His-rich proteins (Fragment) OS=*Plasmodium falciparum* (isolate fcm17 / Senegal) OX=5845 PE=4 SV=1
>Organism: *Plasmodium falciparum (isolate fcm17 / Senegal)*

DPYKERIKSDIRQINESQYLKSLAYKYISGEDYTQYLLLNEVLKDDQDYCTCTRRTIYEE 60

SMDNTVEFAKKMYELSA 77

>S/N:7
>Identifier: sp|P14588|YDH2_PLAFS Uncharacterized protein 3' to Asp-rich and His-rich proteins (Fragment) OS=*Plasmodium falciparum* (isolate fcm17 / Senegal) OX=5845 PE=4 SV=1
>Organism: *Plasmodium falciparum (isolate fcm17 / Senegal)*

MVLVTCNRALAQGDFCLLALIFCHQTCRTPEKHKASQSSAKLVSINISLITSHHRLRHPR 60

RRQHHHRNNFAPTNWYWG 78

>S/N:8
>Identifier: sp|P14589|YDH3_PLAFS Uncharacterized protein 3' to Asp-rich and His-rich proteins (Fragment) OS=*Plasmodium falciparum* (isolate fcm17 / Senegal) OX=5845 PE=4 SV=1
>Organism: *Plasmodium falciparum (isolate fcm17 / Senegal)*

PQYQFVGAKLFRWWCWRRRGWRRRWWLVIKLMLIETSFALDCEALCFSGVRQV 53

**Unreviewed sequences**

>S/N:1a

>Identifier: tr|Q8IDG8|Q8IDG8_PLAF7 Membrane associated histidine-rich protein 2 OS=*Plasmodium falciparum* (isolate 3D7) OX=36329 GN=PF3D7_1353200 PE=4 SV=1
>Organism: *Plasmodium falciparum (isolate 3D7)*

```
MQPCPYDVYNQINHVGTHWAQHLGEHLHHLAHMHQHTPHVHHHIPHVHTLAHDRPCMPIT     60
AFFCRHHEHCSSHLMLIFLLLAFFLVVVYRLYNEVVNSAKTVRIVNITPVNEEHKAEASK     120
EQSKSTSDSSTSTQQTL  137
```

>S/N:1b
>Identifier: tr|Q3ZJL7|Q3ZJL7_PLAFA Histidine-rich protein 2 (Fragment) OS=*Plasmodium falciparum* OX=5833 GN=HRPII PE=4 SV=1
>Organism: *Plasmodium falciparum*

```
AHHAHHVADAHHAHHVADAHHAHHVADAHHAHHVADAHHAHHVADAHHAHHAADAHHAHH     60
AADAHHAHHAADAHHAHHAADAHHAHHAADAHHAHHAADAHHAHHAADAHHAHHAADAHH     120
AHHAAYAHHAHHASDAHHAADAHHAAYAHHAHHAADAHHAADAHHATDAHHAHHAADAHH     180
ATDAHHAHHAADAHHAADAHHAADAHHATDAHHAADAHHAADAHHATDAHHAHHAADAHH     240
AAAHHATDAHHAAAHHEAATH   261
```

>S/N:1c
>Identifier: tr|Q3ZJH6|Q3ZJH6_PLAFA Histidine-rich protein 2 (Fragment) OS=*Plasmodium falciparum* OX=5833 GN=HRPII PE=4 SV=1
>Organism: *Plasmodium falciparum*

```
AHHAHHVADAHHAHHVADAHHAHHAADAHHAHHAAYAHHAHHAADAHHATDAHHAADAHH     60
AADAHHAADAHHAADAHHAHHAADAHHAHHAADAHHAHHAADAHHAHHAADAHHAHHAAD     120
AHHAHHAADAHHAHHAADAHHAHHAADAHHAHHAAYAHHAHHASDAHHAADAHHAAYAHH     180
AHHAADAHHAADAHHATDAHHATDAHHAADAHHAADAHHATDAHHAHHAADAHHAAAHHA     240
TDAHHAAAHHEAATH    255
```

>S/N:1d
>Identifier: tr|Q3ZJJ7|Q3ZJJ7_PLAFA Histidine-rich protein 2 (Fragment) OS=*Plasmodium falciparum* OX=5833 GN=HRPII PE=4 SV=1
>Organism: *Plasmodium falciparum*

```
AHHAHHVADAHHAHHVADAHHAHHVADAHHAHHVADAHHAHHAADAHHAHHAADAHHAHH     60
AADAHHAHHAAYAHHAHHAADAHHAHHAHHAADAHHAHHAADAHHAHHAADAHHAHHAAD     120
AHHAHHAADAHHAHHAADAHHAHHAADAHHAADAHHAADAHHASDAHHAADAHHAAYAHH     180
AHHAADAHHAADAHHATDAHHAADAHHAADAHHATDAHHAHHAADAHHAAAHHATDAHHA     240
AAHHATDAHHAAAHHEAATH    260
```

>S/N:1e
>Identifier: tr|A0A2H4V100|A0A2H4V100_PLAFA Histidine-rich protein 2 (Fragment) OS=*Plasmodium falciparum* OX=5833 GN=HRP2 PE=4 SV=1

>Organism: *Plasmodium falciparum*

CSKNAKGLNLNKRLLHETQAHVDDAHHAHHVADAHHAHHVADAHHAHHAADAHHAHHVAD 60

AHHAHHVADAHHAHHAADAHHAHHAADAHHAHHAHHAADAHHAHHAADAHHAHHAADAHH 120

AHHAADAHHAHHAADAHHAHHAADAHHAHHAADAHHAHHAAYAHHAHHASDAHHAADAHH 180

AAYAHHAHHAADAHHAADAHHATDAHHAADAHHAADAHHATDAHHAADAHHAADAHHAAD 240

AHHAAAHHATDAHHAAAHHATDAHHATDAHHAAAHHEAATHCLRH 285

>S/N:1f
>Identifier: tr|Q3ZJH7|Q3ZJH7_PLAFA Histidine-rich protein 2 (Fragment) OS=*Plasmodium falciparum* OX=5833 GN=HRPII PE=4 SV=1
>Organism: *Plasmodium falciparum*

AHHAHHVADAHHAHHVADAHHAHHVADAHHAHHVADAHHAHHAADAHHAHHAAYAHHAHH 60

AADAHHATDAHHAADAHHAADAHHAADAHHAHHAADAHHAHHAADAHHAHHAADAHHAHH 120

AADAHHAHHAADAHHAHHAAYAHHAHHASDAHHAHHASDAHHAHHAADAHHAAYAHHAHH 180

AADAHHAADAHHATDAHHAADAHHAADAHHATDAHHAADAHHAADAHHAAAHHATDAHHA 240

AAHHATDAHHAAAHHATDAHHAAAHHEAATH 271

>S/N:1g
>Identifier: tr|Q3ZJK0|Q3ZJK0_PLAFA Histidine-rich protein 2 (Fragment) OS=*Plasmodium falciparum* OX=5833 GN=HRPII PE=4 SV=1
>Organism: *Plasmodium falciparum*

AHHAHHVADAHHAHHVADAHHAHHVADAHHAHHVADAHHAHHAADAHHAHHAADAHHAHH 60

AADAHHAHHAADAHHAHHAADAHHAHHAAYAHHAHHAADAHHAHHAHHAADAHHAHHAAD 120

AHHAHHAADAHHAHHAADAHHAHHAADAHHAHHAADAHHAHHAADAHHAHHASDAHHAAD 180

AHHAAYAHHAHHAADAHHAADAHHATDAHHAHHAADAHHATDAHHAADAHHATDAHHAAD 240

AHHAADAHHATDAHHAHHAADAHHAAAHHATDAHHATDAHHAAAHHEAATH 291

>S/N:1h
>Identifier: tr|Q3ZJM1|Q3ZJM1_PLAFA Histidine-rich protein 2 (Fragment) OS=*Plasmodium falciparum* OX=5833 GN=HRPII PE=4 SV=1
>Organism: *Plasmodium falciparum*

AHHAHHVADAHHAHHVADAHHAHHVADAHHAHHAADAHHAHHAADAHHAHHAAYAHHAHH 60

AADAHHATDAHHAADAHHAADAHHAADAHHAADAHHAHHAADAHHAHHAADAHHAHHAAD 120

AHHAHHAADAHHAHHAADAHHAHHAADAHHAHHAADAHHAHHAADAHHAHHAAYAHHAHH 180

AADAHHAAYAHHAHHAADAHHAADAHHATDAHHAADAHHAADAHHAAAHHEAATH 235

>S/N:1i

>Identifier: tr|Q3ZJK8|Q3ZJK8_PLAFA Histidine-rich protein 2 (Fragment) OS=*Plasmodium falciparum* OX=5833 GN=HRPII PE=4 SV=1

>Organism: *Plasmodium falciparum*

AHHAHHVADAHHAHHVADAHHAHHVADAHHAHHVADAHHAHHVADAHHAHHAADAHHAHH 60

AADAHHAHHAADAHHATDAHHAADAHHAADAHHAHHAADAHHAHHAADAHHAHHAADAHH 120

AHHAADAHHAHHAADAHHAHHAADAHHAHHAADAHHAHHAAYAHHAHHASDAHHAADAHH 180

AAYAHHAHHAADAHHAADAHHAADAHHAADAHHATDAHHAHHAADAHHAADAHHAADAHH 240

AAAHHATDAHHAAAHHEAATH 261

>S/N:1j
>Identifier: tr|Q3ZJI4|Q3ZJI4_PLAFA Histidine-rich protein 2 (Fragment) OS=*Plasmodium falciparum* OX=5833 GN=HRPII PE=4 SV=1
>Organism: *Plasmodium falciparum*

AHHAHHVADAHHAHHVADAHHAHHVADAHHAHHAADAHHAHHAADAHHAHHAAYAHHAHH 60

AADAHHATDAHHAADAHHAADAHHAADAHHAADAHHAHHAADAHHAHHAADAHHAHHAAD 120

AHHAHHAADAHHAHHAADAHHAHHAADAHHAHHAADAHHAHHAAYAHHAHHASDAHHAAD 180

AHHAAYAHHAHHAADAHHAADAHHAADAHHATDAHHAADAHHAADAHHAADAHHATDAHH 240

AHHAADAHHAAAHHATDAHHAAAHHEAATH 270

>S/N:1k
>Identifier: tr|Q3ZJM7|Q3ZJM7_PLAFA Histidine-rich protein 2 (Fragment) OS=*Plasmodium falciparum* OX=5833 GN=HRPII PE=4 SV=1
>Organism: *Plasmodium falciparum*

AHHAHHVADAHHAHHVADAHHAHHAADAHHAHHAHHAADAHHAHHAHHAADAHHAHHAAD 60

AHHAHHAADAHHAHHAADAHHAHHAADAHHAHHAADAHHAHHAADAHHAHHAAYAHHAHH 120

ASDAHHAADAHHAAYAHHAHHAADAHHAADAHHATDAHHAHHAADAHHATDAHHAHHAAD 180

AHHATDAHHAHHAADAHHATDAHHAHHAADAHHATDAHHAHHAADAHHAAAHHATDAHHA 240

AAHHATDAHHAAAHHEAATH 260

>S/N:22
>Identifier: CZT62760.1 histidine-rich protein II [*Plasmodium falciparum* 3D7]
>Organism: *Plasmodium falciparum 3D7*

MVSFSKNKVLSAAVFASVLLLDNNNSAFNNNLCSKNAKGLNLNKRLLHETQAHVDDAHHA 60

HHVADAHHAHHVADAHHAHHVADAHHAHHAADAHHAHHAADAHHAHHAADAHHAHHAHHA 120

ADAHHAHHAADAHHAHHAADAHHAHHAADAHHAHHAADAHHAHHASDAHHAHHAAYAHHA 180

HHASDAHHAADAHHAAYAHHAHHAADAHHAADAHHATDAHHAADAHHAADAHHAADAHHA 240

TDAHHAHHAADAHHATDAHHAHHAADAHHAAAHHATDAHHAAAHHATDAHHAAAHHEAAT 300

HCLRH 305

>S/N:23

>Identifier: XP_002808743.1 histidine-rich protein II [*Plasmodium falciparum* 3D7]
>Organism: *Plasmodium falciparum 3D7*

MVSFSKNKVLSAAVFASVLLLDNNNSAFNNNLCSKNAKGLNLNKRLLHETQAHVDDAHHA 60

HHVADAHHAHHVADAHHAHHVADAHHAHHAADAHHAHHAADAHHAHHAADAHHAHHAHHA 120

ADAHHAHHAADAHHAHHAADAHHAHHAADAHHAHHAADAHHAHHASDAHHAHHAAYAHHA 180

HHA**S**DAHHAADAHHAAYAHHAHHAADAHHAADAHHATDAHHAADAHHAADAHHAADAHHA 240

TDAHHAHHAADAHHATDAHHAHHAADAHHAAAHHATDAHHAAAHHATDAHHAAAHHEAAT 300

HCLRH 305

>S/N:24
>Identifier: KAF4330424.1 histidine-rich protein II [*Plasmodium falciparum* NF54]
>Organism: *Plasmodium falciparum NF54*

MVSFSKNKVLSAAVFASVLLLDNNNSAFNNNLCSKNAKGLNLNKRLLHETQAHVDDAHHA 60

HHVADAHHAHHVADAHHAHHVADAHHAHHAADAHHAHHAADAHHAHHAADAHHAHHAHHA 120

ADAHHAHHAADAHHAHHAADAHHAHHAADAHHAHHAADAHHAHHASDAHHAHHAAYAHHA 180

HHASDAHHAADAHHAAYAHHAHHAADAHHAADAHHATDAHHAADAHHAADAHHAADAHHA 240

TDAHHAHHAADAHHATDAHHAHHAADAHHAAAHHATDAHHAAAHHATDAHHAAAHHEAAT 300

HCLRH 305

>S/N:25
>Identifier: PKC46208.1 histidine-rich protein II [*Plasmodium falciparum* NF54]
>Organism: *Plasmodium falciparum NF54*

MVSFSKNKVLSAAVFASVLLLDNNNSAFNNNLCSKNAKGLNLNKRLLHETQAHVDDAHHA 60

HHVADAHHAHHVADAHHAHHVADAHHAHHAADAHHAHHAADAHHAHHAADAHHAHHAHHA 120

ADAHHAHHAADAHHAHHAADAHHAHHAADAHHAHHAADAHHAHHASDAHHAHHAAYAHHA 180

HHASDAHHAADAHHAAYAHHAHHAADAHHAADAHHATDAHHAADAHHAADAHHAADAHHA 240

TDAHHAHHAADAHHATDAHHAHHAADAHHAAAHHATDAHHAAAHHATDAHHAAAHHEAAT 300

HCLRH 305
