## Supplemental file B (Table 1) for "Dynamic programming model for pattern recognition on the *Pf*HRP2 sequence variants: a smart approach to improve malaria immunodiagnostics"

Table 1. Summary of epitope frequency among *Plasmodium falciparum* isolates.

| **S/N** | **Sequence identifier** | **Epitope** | **Epitope frequency** | **Range** |
|---|---|---|---|---|
| 1 | sp\|P09346\|KNOB_PLAFG Knob-associated histidine-rich protein OS=*Plasmodium falciparum* (isolate FCR-3 / Gambia) OX=5838 PE=2 SV=1: / *Plasmodium falciparum* (isolate FCR-3 / Gambia) | | | |
| | | AAYAHHAHHAAY | 0 | Nil |
| | | AHHAADAHH | 0 | Nil |
| | | AHHAADAHHA | 0 | Nil |
| | | AHHAHHA | 0 | Nil |
| | | AHHAHHV | 0 | Nil |
| | | AHHASDAHH | 0 | Nil |
| | | AYAHHAHHAAY | 0 | Nil |
| | | DAHHAADAHH | 0 | Nil |
| | | DAHHAADAHHA | 0 | Nil |
| | | DAHHAHHA | 0 | Nil |
| | | DAHHAHHV | 0 | Nil |
| | | DAHHVADAHH | 0 | Nil |
| | | HAHHAHHAADAHH | 0 | Nil |
| | | HATDAHH | 0 | Nil |
| | | HATDAHHAAD | 0 | Nil |
| | | TDAHHAADAHHAADA | 0 | Nil |
| | | YAHHAHHA | 0 | Nil |
| 2 | sp\|P04930\|HRP_PLAFF Small histidine-alanine-rich protein OS=*Plasmodium falciparum* (isolate FC27 / Papua New Guinea) OX=5837 PE=2 SV=1 *Plasmodium falciparum* (isolate FC27 / Papua New Guinea) | | | |
| | | AAYAHHAHHAAY | 0 | Nil |
| | | AHHAADAHH | 0 | Nil |
| | | AHHAADAHHA | 0 | Nil |
| | | AHHAHHA | 1 | 65 to 71 |
| | | AHHAHHV | 1 | 56 to 62 |
| | | AHHASDAHH | 0 | Nil |
| | | AYAHHAHHAAY | 0 | Nil |
| | | DAHHAADAHH | 0 | Nil |
| | | DAHHAADAHHA | 0 | Nil |
| | | DAHHAHHA | 1 | 64 to 71 |
| | | DAHHAHHV | 1 | 55 to 62 |
| | | DAHHVADAHH | 0 | Nil |
| | | HAHHAHHAADAHH | 0 | Nil |
| | | HATDAHH | 0 | Nil |
| | | HATDAHHAAD | 0 | Nil |
| | | TDAHHAADAHHAADA | 0 | Nil |
| | | YAHHAHHA | 0 | Nil |

| | | | | |
|---|---|---|---|---|
| 3 | sp\|P05227\|HRP1_PLAFA Histidine-rich protein PFHRP-II OS=*Plasmodium falciparum* OX=5833 PE=2 SV=1 *Plasmodium falciparum* | | | |
| | | AAYAHHAHHAAY | 0 | Nil |
| | | AHHAADAHH | 17 | 71 to 79 |
| | | AHHAADAHHA | 13 | 71 to 80 |
| | | AHHAHHA | 14 | 65 to 71 |
| | | AHHAHHV | 1 | 56 to 62 |
| | | AHHASDAHH | 1 | 161 to 169 |
| | | AYAHHAHHAAY | 0 | Nil |
| | | DAHHAADAHH | 6 | 166 to 175 |
| | | DAHHAADAHHA | 6 | 166 to 176 |
| | | DAHHAHHA | 11 | 64 to 71 |
| | | DAHHAHHV | 1 | 55 to 62 |
| | | DAHHVADAHH | 0 | Nil |
| | | HAHHAHHAADAHH | 1 | 67 to 79 |
| | | HATDAHH | 6 | 217 to 223 |
| | | HATDAHHAAD | 3 | 232 to 241 |
| | | TDAHHAADAHHAADA | 2 | 234 to 248 |
| | | YAHHAHHA | 3 | 157 to 164 |
| 4 | sp\|P05228\|HRP2_PLAFA Histidine-rich protein PFHRP-III OS=*Plasmodium falciparum* OX=5833 PE=2 SV=1 *Plasmodium falciparum* | | | |
| | | AAYAHHAHHAAY | 0 | Nil |
| | | AHHAADAHH | 0 | Nil |
| | | AHHAADAHHA | 0 | Nil |
| | | AHHAHHA | 1 | |
| | | AHHAHHV | 2 | |
| | | AHHASDAHH | 0 | Nil |
| | | AYAHHAHHAAY | 0 | Nil |
| | | DAHHAADAHH | 0 | Nil |
| | | DAHHAADAHHA | 0 | Nil |
| | | DAHHAHHA | 1 | |
| | | DAHHAHHV | 2 | |
| | | DAHHVADAHH | 0 | Nil |
| | | HAHHAHHAADAHH | 0 | Nil |
| | | HATDAHH | 0 | Nil |
| | | HATDAHHAAD | 0 | Nil |
| | | TDAHHAADAHHAADA | 0 | Nil |
| | | YAHHAHHA | 0 | Nil |
| 5 | sp\|P14586\|HRP3_PLAFS Histidine-rich protein OS=*Plasmodium falciparum* (isolate fcm17 / Senegal) OX=5845 PE=4 SV=1 *Plasmodium falciparum* (isolate fcm17 / Senegal) | | | |
| | | AAYAHHAHHAAY | 0 | Nil |

| | | | | |
|---|---|---|---|---|
| | | AHHAADAHH | 0 | Nil |
| | | AHHAADAHHA | 0 | Nil |
| | | AHHAHHA | 0 | Nil |
| | | AHHAHHV | 0 | Nil |
| | | AHHASDAHH | 0 | Nil |
| | | AYAHHAHHAAY | 0 | Nil |
| | | DAHHAADAHH | 0 | Nil |
| | | DAHHAADAHHA | 0 | Nil |
| | | DAHHAHHA | 0 | Nil |
| | | DAHHAHHV | 0 | Nil |
| | | DAHHVADAHH | 0 | Nil |
| | | HAHHAHHAADAHH | 0 | Nil |
| | | HATDAHH | 0 | Nil |
| | | HATDAHHAAD | 0 | Nil |
| | | TDAHHAADAHHAADA | 0 | Nil |
| | | YAHHAHHA | 0 | Nil |
| 6 | sp\|P14587\|YDH1_PLAFS Uncharacterized protein 5' to Asp-rich and His-rich proteins (Fragment) OS=*Plasmodium falciparum* (isolate fcm17 / Senegal) OX=5845 PE=4 SV=1 *Plasmodium falciparum* (isolate fcm17 / Senegal) | | | |
| | | AAYAHHAHHAAY | 0 | Nil |
| | | AHHAADAHH | 0 | Nil |
| | | AHHAADAHHA | 0 | Nil |
| | | AHHAHHA | 0 | Nil |
| | | AHHAHHV | 0 | Nil |
| | | AHHASDAHH | 0 | Nil |
| | | AYAHHAHHAAY | 0 | Nil |
| | | DAHHAADAHH | 0 | Nil |
| | | DAHHAADAHHA | 0 | Nil |
| | | DAHHAHHA | 0 | Nil |
| | | DAHHAHHV | 0 | Nil |
| | | DAHHVADAHH | 0 | Nil |
| | | HAHHAHHAADAHH | 0 | Nil |
| | | HATDAHH | 0 | Nil |
| | | HATDAHHAAD | 0 | Nil |
| | | TDAHHAADAHHAADA | 0 | Nil |
| | | YAHHAHHA | 0 | Nil |
| 7 | sp\|P14588\|YDH2_PLAFS Uncharacterized protein 3' to Asp-rich and His-rich proteins (Fragment) OS=*Plasmodium falciparum* (isolate fcm17 / Senegal) OX=5845 PE=4 SV=1 *Plasmodium falciparum* (isolate fcm17 / Senegal) | | | |
| | | AAYAHHAHHAAY | 0 | Nil |
| | | AHHAADAHH | 0 | Nil |
| | | AHHAADAHHA | 0 | Nil |
| | | AHHAHHA | 0 | Nil |

| | | | | |
|---|---|---|---|---|
| | | AHHAHHV | 0 | Nil |
| | | AHHASDAHH | 0 | Nil |
| | | AYAHHAHHAAY | 0 | Nil |
| | | DAHHAADAHH | 0 | Nil |
| | | DAHHAADAHHA | 0 | Nil |
| | | DAHHAHHA | 0 | Nil |
| | | DAHHAHHV | 0 | Nil |
| | | DAHHVADAHH | 0 | Nil |
| | | HAHHAHHAADAHH | 0 | Nil |
| | | HATDAHH | 0 | Nil |
| | | HATDAHHAAD | 0 | Nil |
| | | TDAHHAADAHHAADA | 0 | Nil |
| | | YAHHAHHA | 0 | Nil |
| 8 | sp\|P14589\|YDH3_PLAFS Uncharacterized protein 3' to Asp-rich and His-rich proteins (Fragment) OS=*Plasmodium falciparum* (isolate fcm17 / Senegal) OX=5845 PE=4 SV=1 *Plasmodium falciparum* (isolate fcm17 / Senegal) | | | |
| | | AAYAHHAHHAAY | 0 | Nil |
| | | AHHAADAHH | 0 | Nil |
| | | AHHAADAHHA | 0 | Nil |
| | | AHHAHHA | 0 | Nil |
| | | AHHAHHV | 0 | Nil |
| | | AHHASDAHH | 0 | Nil |
| | | AYAHHAHHAAY | 0 | Nil |
| | | DAHHAADAHH | 0 | Nil |
| | | DAHHAADAHHA | 0 | Nil |
| | | DAHHAHHA | 0 | Nil |
| | | DAHHAHHV | 0 | Nil |
| | | DAHHVADAHH | 0 | Nil |
| | | HAHHAHHAADAHH | 0 | Nil |
| | | HATDAHH | 0 | Nil |
| | | HATDAHHAAD | 0 | Nil |
| | | TDAHHAADAHHAADA | 0 | Nil |
| | | YAHHAHHA | 0 | Nil |
| 1a | tr\|Q8IDG8\|Q8IDG8_PLAF7 Membrane associated histidine-rich protein 2 OS=*Plasmodium falciparum* (isolate 3D7) OX=36329 GN=PF3D7_1353200 PE=4 SV=1 / *Plasmodium falciparum* (isolate 3D7) | | | |
| | | AAYAHHAHHAAY | 0 | Nil |
| | | AHHAADAHH | 0 | Nil |
| | | AHHAADAHHA | 0 | Nil |
| | | AHHAHHA | 0 | Nil |
| | | AHHAHHV | 0 | Nil |
| | | AHHASDAHH | 0 | Nil |
| | | AYAHHAHHAAY | 0 | Nil |

| | | | | |
|---|---|---|---|---|
| | | DAHHAADAHH | 0 | Nil |
| | | DAHHAADAHHA | 0 | Nil |
| | | DAHHAHHA | 0 | Nil |
| | | DAHHAHHV | 0 | Nil |
| | | DAHHVADAHH | 0 | Nil |
| | | HAHHAHHAADAHH | 0 | Nil |
| | | HATDAHH | 0 | Nil |
| | | HATDAHHAAD | 0 | Nil |
| | | TDAHHAADAHHAADA | 0 | Nil |
| | | YAHHAHHA | 0 | Nil |
| 1b | tr\|Q3ZJL7\|Q3ZJL7_PLAFA Histidine-rich protein 2 (Fragment) OS=*Plasmodium falciparum* OX=5833 GN=HRPII PE=4 SV=1 / *Plasmodium falciparum* | | | |
| | | AAYAHHAHHAAY | 0 | Nil |
| | | AHHAADAHH | 15 | |
| | | AHHAADAHHA | 11 | |
| | | AHHAHHA | 14 | |
| | | AHHAHHV | 5 | |
| | | AHHASDAHH | 1 | |
| | | AYAHHAHHAAY | 0 | Nil |
| | | DAHHAADAHH | 4 | |
| | | DAHHAADAHHA | 4 | |
| | | DAHHAHHA | 12 | |
| | | DAHHAHHV | 4 | |
| | | DAHHVADAHH | 0 | Nil |
| | | HAHHAHHAADAHH | 0 | Nil |
| | | HATDAHH | 5 | |
| | | HATDAHHAAD | 1 | |
| | | TDAHHAADAHHAADA | 1 | |
| | | YAHHAHHA | 2 | |
| 1c | tr\|Q3ZJH6\|Q3ZJH6_PLAFA Histidine-rich protein 2 (Fragment) OS=*Plasmodium falciparum* OX=5833 GN=HRPII PE=4 SV=1 / *Plasmodium falciparum* | | | |
| | | AAYAHHAHHAAY | 0 | Nil |
| | | AHHAADAHH | 16 | |
| | | AHHAADAHHA | 12 | |
| | | AHHAHHA | 15 | |
| | | AHHAHHV | 2 | |
| | | AHHASDAHH | 1 | |
| | | AYAHHAHHAAY | 0 | Nil |
| | | DAHHAADAHH | 5 | |
| | | DAHHAADAHHA | 5 | |
| | | DAHHAHHA | 12 | |
| | | DAHHAHHV | 1 | |
| | | DAHHVADAHH | 0 | Nil |

| | | | | |
|---|---|---|---|---|
| | | HAHHAHHAADAHH | 0 | Nil |
| | | HATDAHH | 4 | |
| | | HATDAHHAAD | 2 | |
| | | TDAHHAADAHHAADA | 2 | |
| | | YAHHAHHA | 3 | |
| 1d | : tr\|Q3ZJJ7\|Q3ZJJ7_PLAFA Histidine-rich protein 2 (Fragment) OS=*Plasmodium falciparum* OX=5833 GN=HRPII PE=4 SV=1 / *Plasmodium falciparum* | | | |
| | | AAYAHHAHHAAY | 0 | Nil |
| | | AHHAADAHH | 16 | |
| | | AHHAADAHHA | 12 | |
| | | AHHAHHA | 14 | |
| | | AHHAHHV | 4 | |
| | | AHHASDAHH | 1 | |
| | | AYAHHAHHAAY | 0 | Nil |
| | | DAHHAADAHH | 4 | |
| | | DAHHAADAHHA | 4 | |
| | | DAHHAHHA | 12 | |
| | | DAHHAHHV | 3 | |
| | | DAHHVADAHH | 0 | Nil |
| | | HAHHAHHAADAHH | 1 | |
| | | HATDAHH | 4 | |
| | | HATDAHHAAD | 1 | |
| | | TDAHHAADAHHAADA | 1 | |
| | | YAHHAHHA | 2 | |
| 1e | tr\|A0A2H4V100\|A0A2H4V100_PLAFA Histidine-rich protein 2 (Fragment) OS=*Plasmodium falciparum* OX=5833 GN=HRP2 PE=4 SV=1 / *Plasmodium falciparum* | | | |
| | | AAYAHHAHHAAY | 0 | Nil |
| | | AHHAADAHH | 15 | |
| | | AHHAADAHHA | 11 | |
| | | AHHAHHA | 13 | |
| | | AHHAHHV | 4 | |
| | | AHHASDAHH | 1 | |
| | | AYAHHAHHAAY | 0 | |
| | | DAHHAADAHH | 5 | |
| | | DAHHAADAHHA | 5 | |
| | | DAHHAHHA | 11 | |
| | | DAHHAHHV | 4 | |
| | | DAHHVADAHH | 0 | Nil |
| | | HAHHAHHAADAHH | 1 | |
| | | HATDAHH | 4 | |
| | | HATDAHHAAD | 2 | |
| | | TDAHHAADAHHAADA | 2 | |

| | | | | |
|---|---|---|---|---|
| | | YAHHAHHA | 2 | |
| 1f | tr\|Q3ZJH7\|Q3ZJH7_PLAFA Histidine-rich protein 2 (Fragment) OS=*Plasmodium falciparum* OX=5833 GN=HRPII PE=4 SV=1 / *Plasmodium falciparum* | | | |
| | | AAYAHHAHHAAY | 0 | Nil |
| | | AHHAADAHH | 13 | |
| | | AHHAADAHHA | 10 | |
| | | AHHAHHA | 13 | |
| | | AHHAHHV | 4 | |
| | | AHHASDAHH | 2 | |
| | | AYAHHAHHAAY | 0 | Nil |
| | | DAHHAADAHH | 5 | |
| | | DAHHAADAHHA | 5 | |
| | | DAHHAHHA | 10 | |
| | | DAHHAHHV | 3 | |
| | | DAHHVADAHH | 0 | Nil |
| | | HAHHAHHAADAHH | 0 | Nil |
| | | HATDAHH | 6 | |
| | | HATDAHHAAD | 3 | |
| | | TDAHHAADAHHAADA | 3 | |
| | | YAHHAHHA | 3 | |
| 1g | tr\|Q3ZJK0\|Q3ZJK0_PLAFA Histidine-rich protein 2 (Fragment) OS=*Plasmodium falciparum* OX=5833 GN=HRPII PE=4 SV=1 / *Plasmodium falciparum* | | | |
| | | AAYAHHAHHAAY | 0 | Nil |
| | | AHHAADAHH | 19 | |
| | | AHHAADAHHA | 14 | |
| | | AHHAHHA | 18 | |
| | | AHHAHHV | 4 | |
| | | AHHASDAHH | 1 | |
| | | AYAHHAHHAAY | 0 | Nil |
| | | DAHHAADAHH | 4 | |
| | | DAHHAADAHHA | 4 | |
| | | DAHHAHHA | 16 | |
| | | DAHHAHHV | 3 | |
| | | DAHHVADAHH | 0 | Nil |
| | | HAHHAHHAADAHH | 1 | |
| | | HATDAHH | 5 | |
| | | HATDAHHAAD | 2 | |
| | | TDAHHAADAHHAADA | 1 | |
| | | YAHHAHHA | 2 | |
| 1h | tr\|Q3ZJM1\|Q3ZJM1_PLAFA Histidine-rich protein 2 (Fragment) OS=*Plasmodium falciparum* OX=5833 GN=HRPII PE=4 SV=1 / *Plasmodium falciparum* | | | |
| | | AAYAHHAHHAAY | 0 | Nil |

| | | | | |
|---|---|---|---|---|
| | | AHHAADAHH | 16 | |
| | | AHHAADAHHA | 11 | |
| | | AHHAHHA | 15 | |
| | | AHHAHHV | 3 | |
| | | AHHASDAHH | 0 | Nil |
| | | AYAHHAHHAAY | 0 | Nil |
| | | DAHHAADAHH | 4 | |
| | | DAHHAADAHHA | 4 | |
| | | DAHHAHHA | 12 | |
| | | DAHHAHHV | 2 | |
| | | DAHHVADAHH | 0 | Nil |
| | | HAHHAHHAADAHH | 0 | Nil |
| | | HATDAHH | 2 | |
| | | HATDAHHAAD | 2 | |
| | | TDAHHAADAHHAADA | 2 | |
| | | YAHHAHHA | 3 | |
| 1i | tr\|Q3ZJK8\|Q3ZJK8_PLAFA Histidine-rich protein 2 (Fragment) OS=*Plasmodium falciparum* OX=5833 GN=HRPII PE=4 SV=1 / *Plasmodium falciparum* | | | |
| | | AAYAHHAHHAAY | 0 | Nil |
| | | AHHAADAHH | 16 | |
| | | AHHAADAHHA | 12 | |
| | | AHHAHHA | 14 | |
| | | AHHAHHV | 5 | |
| | | AHHASDAHH | 1 | |
| | | AYAHHAHHAAY | 0 | Nil |
| | | DAHHAADAHH | 5 | |
| | | DAHHAADAHHA | 5 | |
| | | DAHHAHHA | 12 | |
| | | DAHHAHHV | 4 | |
| | | DAHHVADAHH | 0 | Nil |
| | | HAHHAHHAADAHH | 0 | Nil |
| | | HATDAHH | 3 | |
| | | HATDAHHAAD | 1 | |
| | | TDAHHAADAHHAADA | 1 | |
| | | YAHHAHHA | 2 | |
| 1j | tr\|Q3ZJI4\|Q3ZJI4_PLAFA Histidine-rich protein 2 (Fragment) OS=*Plasmodium falciparum* OX=5833 GN=HRPII PE=4 SV=1 / *Plasmodium falciparum* | | | |
| | | AAYAHHAHHAAY | 0 | Nil |
| | | AHHAADAHH | 18 | |
| | | AHHAADAHHA | 14 | |
| | | AHHAHHA | 15 | |
| | | AHHAHHV | 3 | |
| | | AHHASDAHH | 1 | |

| | | | | |
|---|---|---|---|---|
| | | AYAHHAHHAAY | 0 | Nil |
| | | DAHHAADAHH | 6 | |
| | | DAHHAADAHHA | 6 | |
| | | DAHHAHHA | 12 | |
| | | DAHHAHHV | 2 | |
| | | DAHHVADAHH | 0 | Nil |
| | | HAHHAHHAADAHH | 0 | Nil |
| | | HATDAHH | 4 | |
| | | HATDAHHAAD | 2 | |
| | | TDAHHAADAHHAADA | 2 | |
| | | YAHHAHHA | 3 | |
| 1k | tr\|Q3ZJM7\|Q3ZJM7_PLAFA Histidine-rich protein 2 (Fragment) OS=*Plasmodium falciparum* OX=5833 GN=HRPII PE=4 SV=1 / *Plasmodium falciparum* | | | |
| | | AAYAHHAHHAAY | 0 | Nil |
| | | AHHAADAHH | 16 | |
| | | AHHAADAHHA | 13 | |
| | | AHHAHHA | 17 | |
| | | AHHAHHV | 2 | |
| | | AHHASDAHH | 1 | |
| | | AYAHHAHHAAY | 0 | Nil |
| | | DAHHAADAHH | 2 | |
| | | DAHHAADAHHA | 2 | |
| | | DAHHAHHA | 15 | |
| | | DAHHAHHV | 1 | |
| | | DAHHVADAHH | 0 | Nil |
| | | HAHHAHHAADAHH | 1 | |
| | | HATDAHH | 7 | |
| | | HATDAHHAAD | 0 | Nil |
| | | TDAHHAADAHHAADA | 0 | Nil |
| | | YAHHAHHA | 2 | |
| 22 | CZT62760.1 histidine-rich protein II [*Plasmodium falciparum* 3D7] / *Plasmodium falciparum* 3D7 | | | |
| | | AAYAHHAHHAAY | 0 | Nil |
| | | AHHAADAHH | 14 | |
| | | AHHAADAHHA | 11 | |
| | | AHHAHHA | 14 | |
| | | AHHAHHV | 3 | |
| | | AHHASDAHH | 2 | |
| | | AYAHHAHHAAY | 0 | Nil |
| | | DAHHAADAHH | 4 | |
| | | DAHHAADAHHA | 4 | |
| | | DAHHAHHA | 12 | |
| | | DAHHAHHV | 3 | |

| | | | | |
|---|---|---|---|---|
| | | DAHHVADAHH | 0 | Nil |
| | | HAHHAHHAADAHH | 1 | |
| | | HATDAHH | 5 | |
| | | HATDAHHAAD | 1 | |
| | | TDAHHAADAHHAADA | 1 | |
| | | YAHHAHHA | 2 | |
| 23 | XP_002808743.1 histidine-rich protein II [*Plasmodium falciparum* 3D7] / *Plasmodium falciparum* 3D7 | | | |
| | | AAYAHHAHHAAY | 0 | Nil |
| | | AHHAADAHH | 14 | |
| | | AHHAADAHHA | 11 | |
| | | AHHAHHA | 14 | |
| | | AHHAHHV | 3 | |
| | | AHHASDAHH | 2 | |
| | | AYAHHAHHAAY | 0 | Nil |
| | | DAHHAADAHH | 4 | |
| | | DAHHAADAHHA | 4 | |
| | | DAHHAHHA | 12 | |
| | | DAHHAHHV | 3 | |
| | | DAHHVADAHH | 0 | Nil |
| | | HAHHAHHAADAHH | 1 | |
| | | HATDAHH | 5 | |
| | | HATDAHHAAD | 1 | |
| | | TDAHHAADAHHAADA | 1 | |
| | | YAHHAHHA | 2 | |
| 24 | KAF4330424.1 histidine-rich protein II [*Plasmodium falciparum* NF54] / *Plasmodium falciparum* NF54 | | | |
| | | AAYAHHAHHAAY | 0 | Nil |
| | | AHHAADAHH | 14 | |
| | | AHHAADAHHA | 11 | |
| | | AHHAHHA | 14 | |
| | | AHHAHHV | 3 | |
| | | AHHASDAHH | 2 | |
| | | AYAHHAHHAAY | 0 | Nil |
| | | DAHHAADAHH | 4 | |
| | | DAHHAADAHHA | 4 | |
| | | DAHHAHHA | 12 | |
| | | DAHHAHHV | 3 | |
| | | DAHHVADAHH | 0 | Nil |
| | | HAHHAHHAADAHH | 1 | |
| | | HATDAHH | 5 | |
| | | HATDAHHAAD | 1 | |
| | | TDAHHAADAHHAADA | 1 | |

| | | | | |
|---|---|---|---|---|
| | | YAHHAHHA | 2 | |
| 25 | PKC46208.1 histidine-rich protein II [*Plasmodium falciparum* NF54] / Organism: *Plasmodium falciparum* NF54 | | | |
| | | AAYAHHAHHAAY | 0 | Nil |
| | | AHHAADAHH | 14 | |
| | | AHHAADAHHA | 11 | |
| | | AHHAHHA | 14 | |
| | | AHHAHHV | 3 | |
| | | AHHASDAHH | 2 | |
| | | AYAHHAHHAAY | 0 | Nil |
| | | DAHHAADAHH | 4 | |
| | | DAHHAADAHHA | 4 | |
| | | DAHHAHHA | 12 | |
| | | DAHHAHHV | 3 | |
| | | DAHHVADAHH | 0 | Nil |
| | | HAHHAHHAADAHH | 1 | |
| | | HATDAHH | 5 | |
| | | HATDAHHAAD | 1 | |
| | | TDAHHAADAHHAADA | 1 | |
| | | YAHHAHHA | 2 | |
